# A comparison between a Random Forest model and the Kidney Failure Risk Equation to predict progression to kidney failure

**DOI:** 10.1101/2023.05.16.23290068

**Authors:** Marina Wainstein, Amir Kamel Rahimi, Ivor Katz, Helen Healy, Saiyini Pirabhahar, Kylie Turner, Sally Shrapnel

**Author notes:** **Address for Correspondence and Reprints** Dr Marina Wainstein, NHMRC CKD.CRE, Health Sciences Building, Level 8, Royal Brisbane and Women’s Hospital, Butterfield Street, Herston, Brisbane, Queensland, 4006, Australia.

## Abstract

**Background:** Machine learning may offer a superior alternative to traditional prediction tools when used to model complicated, nonlinear interactions between variables. While modern machine learning methods are tagged as “black boxes”, the random forest (RF) classifier can be interrogated to understand the contribution of input variables (feature importance), thereby improving the interpretability of its predictions. We hypothesized that a random forest (RF) classifier would have equivalent, if not superior, performance to the 4-variable Kidney Failure Risk Equation (KFRE) in predicting progression to end stage kidney disease (ESKD) in a chronic kidney disease (CKD) population and explored the impact of serum creatinine and primary renal disease on prediction accuracy.

**Methods:** A 2-year risk of ESKD was calculated using the 4-variable KFRE and compared to a RF model using the same four variables (age, gender, eGFR and urine albumin creatinine ratio). Four more RF models were developed using a combination of these as well as serum creatinine and primary renal disease. Performance of the KFRE and RF models was assessed by area under a receiver operating (AUC ROC) curve and feature importance was evaluated for each RF model.

**Results:** Of 1365 patients with CKD from two renal units included in the analysis, 208 progressed to ESKD in the 2-year follow-up period. The AUC ROC for KFRE was 0.95 (95% confidence interval, 0.93 – 0.96) and for the RF model using the same 4 variables 0.97. The remaining four RF models had similar performance (AUC ROC 0.97 – 0.98). In the RF models, eGFR and serum creatinine had the largest effect on risk prediction while gender had the smallest.

**Conclusions:** Our findings suggest that RF models provide a potential tool to predict CKD progression with competing accuracy and interpretability to the current benchmark equation. They therefore warrant validation in larger and more diverse populations

## Introduction

Accurate detection and prediction of chronic kidney disease (CKD) trajectory are key to the fundamental clinical questions of timing and selection of effective therapies and management plans. The Kidney Failure Risk Equation (KFRE), developed in a Canadian CKD population and subsequently validated in multiple international cohorts, is generally accepted as the best prediction tool for progression of kidney disease [1-5]. The 4-and 8-variable equations were selected from a range of candidate multivariate regression models designed to predict ESKD using demographic, clinical and laboratory variables in pre-defined time frames from 1 to 5 years. Lower eGFR (estimated glomerular filtration rate ml/min using CKD-EPI equation), higher concentrations of albuminuria (albumin in urine in mg/creatinine), younger age and male gender in the 4-variable model, added to lower serum albumin, serum calcium, serum bicarbonate and higher serum phosphate in the 8 variable model predicted faster progression to ESKD (defined as requiring dialysis, having a renal transplant or reaching an eGFR less than 10 ml/min/1.73^2^)[1]. The KFRE equation has been validated in several multinational cohorts, achieving excellent predictive power (AUC ROC 80-90%)[2, 6], and has been used to inform risk thresholds for clinical management like specialist nephrology referrals, timing of dialysis education and vascular access creation[4, 7, 8].

One important *potential* limitation in the predictive performance of the KFRE is its development using traditional regression techniques which assume a linear relationship between the independent and dependent variables. In the context of a disease like CKD in which trajectory and clinical tempo are subject to a complex, patient-specific interaction between initiating and perpetuating risk factors, such an assumption may have a significant impact on the performance of a prediction tool[9].

Machine learning may offer a superior alternative to traditional statistical prediction models, particularly when used to model complicated, nonlinear interactions between variables and when datasets are large and poorly defined[10, 11]. The use of general-purpose learning algorithms in these models means that a larger number of variables, often not known to contribute to the outcome, can potentially be used to improve predictive accuracy. In nephrology, machine learning models have already been used in different patient cohorts to predict a broad range of outcomes including, among others, acute kidney injury events, transplant graft survival and optimal dialysis targets and mortality[12-16]. Studies using various CKD populations have been able to predict disease progression, defined by eGFR decline or ESKD status, using combinations of supervised classification and regression models, with striking accuracy[17-21].

One specific learning algorithm that is gaining traction in the healthcare context is the random forest (RF). A RF is a non-parametric, classification algorithm derived from an ensemble of individual decision trees and their predictions [22]. Performance of RFs are further improved through bagging or boosting which allows individual trees to be developed on randomly selected subsets of the data (bagging) or iteratively using the entire training dataset and adjusting weighting of cases according to classification errors (boosting)[23]. These techniques have been shown to both improve prediction accuracy and reduce the probability of model overfitting[24]. Additionally, RFs have the capacity to select and rank variables according to their impact on its predictions, conferring additional insight and understanding of the role of individual variables.

A comparison of predictive accuracy between the KFRE and a machine learning approach has not yet been performed. Thus, the objective of this study was to develop a random forest model able to predict progression to ESKD in patients with CKD and compare its performance to the KFRE using the same set of four variables. We hypothesized that, given the non-linear and complex relationship between the variables contributing to CKD progression, the random forest would have equivalent if not superior predictive performance to the KFRE. Furthermore, given the inclusion of age, gender and creatinine in the estimated GFR formula, we hypothesized that replacing eGFR with creatinine in several models would not significantly affect risk prediction accuracy, thereby allowing us to bypass the need to use such a formula that has its own inherent limitations[25]. Finally, we explored the effect of adding patients’ primary renal disease to ESKD risk prediction, noting previous conflicting findings in the literature of the role of this variable in predicting CKD progression[9].

## Methods

### Study Population

Patients were divided into two populations, the “Starter Population” included patients with complete data for age, gender, urine ACR and eGFR used to compute the 4-variable KFRE and develop RF model A, and the second population, named “Derived population”, included patients from the Starter Population who in addition to the four original variables also had available data collected for serum creatinine and primary renal disease and were used to develop RF models B, C, D and E (Fig. 1). Patients were selected from two different kidney health services in Australia, the Royal Brisbane and Women’s hospital within the broader Metro North Hospital and Health Service (MNHHS) in Brisbane and St George and Sutherland Hospitals in the South Eastern Sydney Local Health District (SESLHD). The patients selected from the MNHHS renal unit had previously consented to a state-wide CKD registry, allowing access to their data at consent, from 2011 onwards and information on outcomes which we censored in this study at 2018. St George and Sutherland renal service maintains, since 2010, a database of CKD patients with information relating to demographic information, pathology data and outcomes (including hospitalizations and commencement of dialysis or transplantation) extracted from electronic medical records, pathology providers and clinic letters.

**Fig 1.**
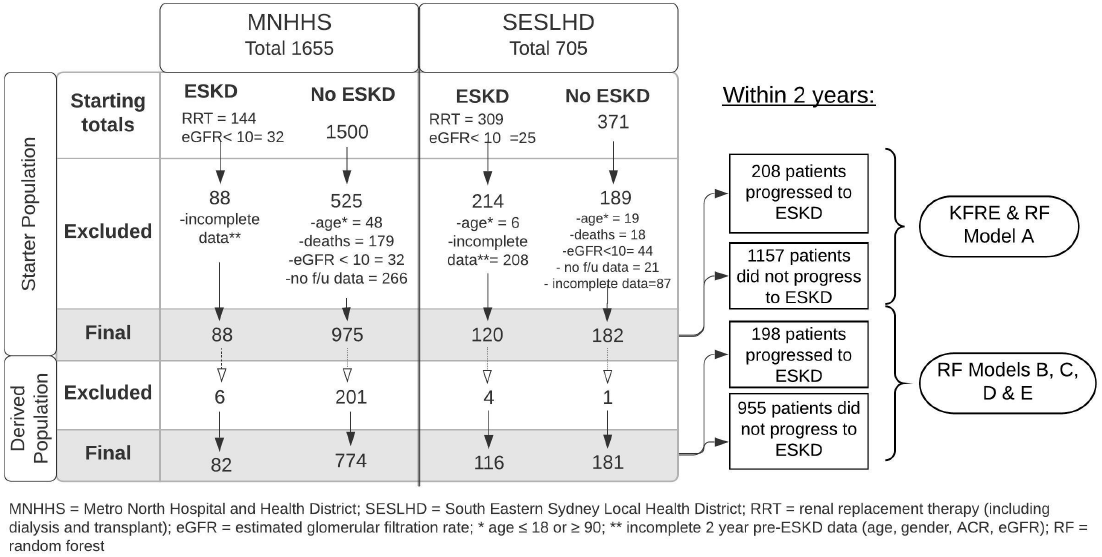
Flowchart of patients used in the final KFRE and RF models in the Starter Population and Derived Population

Patients from both units were selected if their eGFR at time of consent (or entry into the database) was less than 60 ml/min, a selection criterion of the KFRE, and they were between the ages of 18 and 90. End-stage kidney disease (ESKD) was defined as needing to commence dialysis, receiving a renal transplant or reaching an eGFR less than 10 ml/min/1.73m^2^. For the Starter Population, patients that had reached ESKD were identified in both databases (from 2011 to 2017 in MNHHS and from 2010 to 2018 in SESLHD) and information pertaining to the 4 variables were searched retrospectively within the period closest to 2 years preceding the event, ESKD, allowing a period of 3 months before and after this date to record urine ACR and eGFR. Patients with no available data during this period of time were excluded (Fig. 1). Patients were labelled as “No ESKD” if, in the two years after being consented or entered into the database, they had not reached ESKD (as defined above). Patients who met this criterion were further excluded if in this two-year period they either died, required acute dialysis or no follow up eGFR could be obtained at two years to confirm they had not reached ESKD (Fig. 1). The Derived Population was composed of patients from the Starter Population for whom data for serum creatinine and primary renal disease (PRD, ANZDATA coding) could be obtained in the same time period as was originally recorded for urine ACR and eGFR (Fig 1.)

### Ethics Approval

Approval for the use of the SESLHD patient data was granted by Human Research Ethics Committee of the South Eastern Sydney Local Health District (HREC Ref 11/STG/212). The MNHHS and QLD registry study was approved by the Queensland Health Human Research Ethics Committee (HREC approval: HREC/10/QHC/41 in November 2010.

### Statistical Analysis

#### Two-year KFRE scores

Two-year KFRE scores were calculated for all patients using age, gender, eGFR (ml/min using CKD-EPI equation) and urine ACR (mg/g) (KFRE variables) in the two years before reaching ESKD or, in the case of patients not reaching this outcome, within two years (No ESKD) of time of consent into the registry or database.

#### Random Forest model implementation

Five RF models were developed. Model A used the same four variables used to calculate the 4-variable KFRE equation in Starter Population while models B to E were developed using a combination of these variables as well as serum creatinine in the Derived Population (Fig 1 and Table 1). A random forest classifier was implemented using sklearn.ensemble.RandomForestClassifier package[26] with class weighting to address class imbalance. We explored the contribution of individual variables to each risk prediction by using the feature importance ranking feature of the model.

**Table 1.**

| **Main population** |  |  |
| --- | --- | --- |
|  | **ESKD** | **No ESKD** |
| **No. of Participants (% total)** | 208 | 1157 |
| **Age, mean (SD), y** | 66 (15) | 71 (13) |
| **Male gender (% total)** | 130 (63) | 605 (52) |
| **eGFR (ml/min), mean (SD)** | 21 (8) | 37 (11) |
| **Urine ACR (mg/g), median (IQR)** | 795 (1640) | 91 (276) |

#### Performance Metrics

Performance of the KFRE and RF models as predictors of risk of ESKD was assessed by area under a receiver operating (AUC ROC) curve. Sensitivity, specificity, precision and F1 mean were calculated for a KFRE test threshold of 10% and for each RF model. In addition, out-of-bag scores were calculated using sklearn.ensemble for each RF model.

## Results

### Patient characteristics

The Starter Population consisted of 1365 patients, 1063 from MNHHS and 302 from St SESLHD, with 208 reaching ESKD and 1157 not reaching this outcome (Fig 1). 613 patients were excluded from the final analysis from MNHHS and 302 from SESLHD databases. Just over half of all patients in the Starter Population were male with mean age of 70, mean eGFR 35 ml/min/1.73m^2^ and median urine ACR 121 mg/g two years before an ESKD event or at time of consent into the database or registry (table 1). The Derived Population had 1151 patients with similar demographic and laboratory figures as the Starter Population. Additional characteristics included a mean Creatinine of 174 umol/L and a predominance of renal vascular disease (42%) and diabetic nephropathy (27%) as primary renal disease (stable 1).

### Model performance

The area under the receiver operating curve (AUC ROC) for the 2-year KFRE was 95.0 (95% confidence interval, 93 – 96) and model A, which used the same four variables, had an AUC ROC of 97.0 (table 2). Performance of all models was optimized using a 1:3 class weight hyperparameter, which resulted in similar performance for models B, C, D and E (AUC ROC 97.0 – 98.0). Specificity for all models and KFRE was excellent (91 – 94%) while sensitivity was lower for the KFRE (80%) and highest for Model B (92%). The F1 mean was consistently higher for the RF models (92-93% than for the KFRE (73%).

**Table 2.**

|  | **Main Population** |  | **Sub-Population** |  |  |  |
| --- | --- | --- | --- | --- | --- | --- |
|  | 2-year KFRE | Model A | Model B | Model C | Model D | Model E |
| **Variables Used** | Age, gender, eGFR, urine ACR |  | Age, gender, creatinine, urine ACR | Age, gender, eGFR, urine ACR, PRD | Age, gender, creatinine, urine ACR, PRD | Age, gender, eGFR, creatinine, urine ACR, PRD |
| **AUC_ROC** | 95.0 | 97.0 | 97.0 | 98.0 | 97.0 | 98.0 |
| **Sensitivity** | 80.0 | 86.0 | 92.0 | 90.0 | 91.0 | 88.0 |
| **Specificity** | 93.0 | 94.0 | 91.0 | 93.0 | 92.0 | 92.0 |
| **Precision (PPV)** | 68.0 | 72.0 | 69.0 | 73.0 | 70.0 | 71.0 |
| **F1 Mean** | 73.0 | 93.0 | 91.9 | 92.9 | 92.2 | 92.0 |
| **Out-of-bag-score** | - | 90.3 | 89.1 | 90.4 | 89.4 | 89.8 |
| **Feature Importance Weights** |  |  |  |  |  |  |
| Age | - | 6.7 | 6.5 | 6.1 | 4.4 | 3.8 |
| Gender | - | 0.7 | 0.4 | 0.6 | 0.4 | 0.3 |
| eGFR | - | **61.2** | - | **52.4** | - | 27.8 |
| Urine ACR | - | 31.4 | 31.6 | 33.0 | 29.9 | 21.0 |
| Creatinine | - |  | **61.6** | - | **58.3** | **42.1** |
| PRD | - |  | - | 8.0 | 7.1 | 5.1 |

Models A, B, C, D, E are all Random Forest models.
All performance metrics and feature importance weights presented as percentages (%)

#### Variable feature importance

Across all RF models, gender had the smallest impact on risk prediction, followed by age and primary renal disease (all less than 10% weight) (Table 2). Estimated GFR and creatinine had the largest effect on predictions in models that featured either one (Models A to D, weight 52.4 to 61.6%), with urine ACR being a close second (weight 21 - 33%). In model E, which included all six variables, creatinine had the strongest predictive weight (42.1%) followed by eGFR (27.8%) and urine ACR (21.0%).

## Discussion

In this study we developed an RF model that had comparable performance and greater sensitivity to the KRFE in predicting 2-year risk of progression to ESKD using the same four variables. Subsequent RF models displayed equivalent predictive weighting of creatinine and eGFR and little gain in performance with the addition of primary renal disease.

The accuracy in predicting CKD progression using RF models has been reported in several studies[18, 27], but none have compared it directly to a validated risk prediction score. Although we are unable to comment on clinical utility, our findings suggest that RF models can perform just as well as the KFRE and have the added potential benefit of providing information on a wider range of clinical variables, as is found in electronic health records. Further studies are needed to examine the possible contribution of these models to clinical decision-making and patient care.

In rare events like progression to ESKD in CKD patients, specificity is clinically favoured over sensitivity. Both prediction models gave good specificity. RF produced superior sensitivity, giving clinicians confidence to, for example, discharge patients to primary healthcare providers where prediction of progression to ESKD is low. The improved sensitivity of our RF models may be a product of the class weighting that we applied to address the imbalance in the number of patients who reached ESKD versus those that did not. By increasing the weight of the former (and smaller) group in the final predictions, the model may have increased the number of true positive results. This hypothesis, and its clinical impact, would need to be examined in further studies using varying degrees of class imbalance in sample populations.

The interchangeable value of creatinine and eGFR as measures of kidney function and biomarkers of disease progression is reflected in the lack of consensus in existing risk prediction scores [28]. The greater contribution of creatinine over eGFR in Model E may signal that creatinine, which appears in a patient’s routine biochemical profile, is a sufficient measure of renal function and therefore disease progression, without needing to be contextualized into the eGFR equation which includes gender, age and race. This has implications for our current staging and risk-stratification of renal disease frameworks which are eGFR based. It could allow for predictive ML models to be used in low resource settings where point of care creatinine is the best available biomarker of kidney function. The relative merits of both variables in definitions of CKD progression and their comparative contributions in models with a broader range of predictor variables continues to be a gap in the literature.

The marginal improvement in model performance using PRD may be explained by the already high level of performance of the existing models (Models A and B) that is driven mainly by the non-PRD variables of eGFR, serum creatinine or urine ACR. It should be noted that previous studies have not seen a close relationship between PRD and CKD progression[9] which may explain why PRD does not feature in the latest KDIGO staging of CKD[8]. Nevertheless, any conclusion on the role of PRD in our models’ predictions must be interpreted within the context of a heterogeneous CKD population and the potential for resultant training bias. Nevertheless, it should be noted that the PRD profile of our population closely resembles that of the wider Australian population[29].

The small sample size in our study has the potential to limit the model’s learning and ability to generalize to other populations. To address and minimize this we used out-of-bag error for internal validation to avoid splitting our original sample into a training and test set as well as class weighting to improve the models’ learning. In addition, our population consisted of patients from two different states and units in Australia which may result in a more diverse profile. The use of retrospective patient data, particularly in the selection of patients that progressed to ESKD, may have resulted in selection bias and an overestimation of risk in this group.

Our study is the first to validate the KFRE in an Australian cohort and to compare it to a machine learning model, an approach that may become the standard in prediction modelling in a Big Data future. The use of RF models allowed us to interpret the predictions, gaining insights into the role of each predictor variable while performing equivalently to the KFRE in this population. In conclusion, our findings suggest that RF models provide a potential tool to predict CKD progression and therefore warrant validation in larger and more diverse populations.

## Data Availability

All data produced in the present work are contained in the manuscript

## Acknowledgements

This study was not funded

## Conflict of interest statement

All the authors declared no competing interests

The results presented in this paper have not been published previously in whole or part, except in abstract format.

## Summary table

What is already known about this subject:

- The Kidney Failure Risk Equation (KFRE), developed from several candidate multivariate regression models, is the most validated tool used to predict progression to end-stage kidney disease (ESKD).
- A potential limitation in the predictive performance of the KFRE is its development using traditional regression techniques which assume a linear relationship between the independent and dependent variables.
- Machine learning models such as a random forest (RF) may have superior performance when applied to large, poorly defined datasets where no linearity can be assumed between variables while maintaining some prediction interpretability. A comparison between a RF and the KFRE is therefore warranted.

What this study adds:

- We developed a RF with comparable performance and greater sensitivity to the KRFE using the same four variables; subsequent RF models displayed equivalent predictive weighting of creatinine and eGFR and little gain in performance with the addition of primary renal disease
- In the context of a complex disease such as CKD and the potential to widen the scope of predictive variables from electronic health records, validation of machine learning models in more diverse populations is crucial.

What impact this may have on practice or policy:

- The comparable performance of a machine learning model to the current benchmark in CKD risk prediction coupled with the capacity to incorporate a greater number of presumably unrelated variables while providing it’s user with interpretable predictions, may render them the optimal prediction tool.

## Supplementary Material

**sTable 1.**
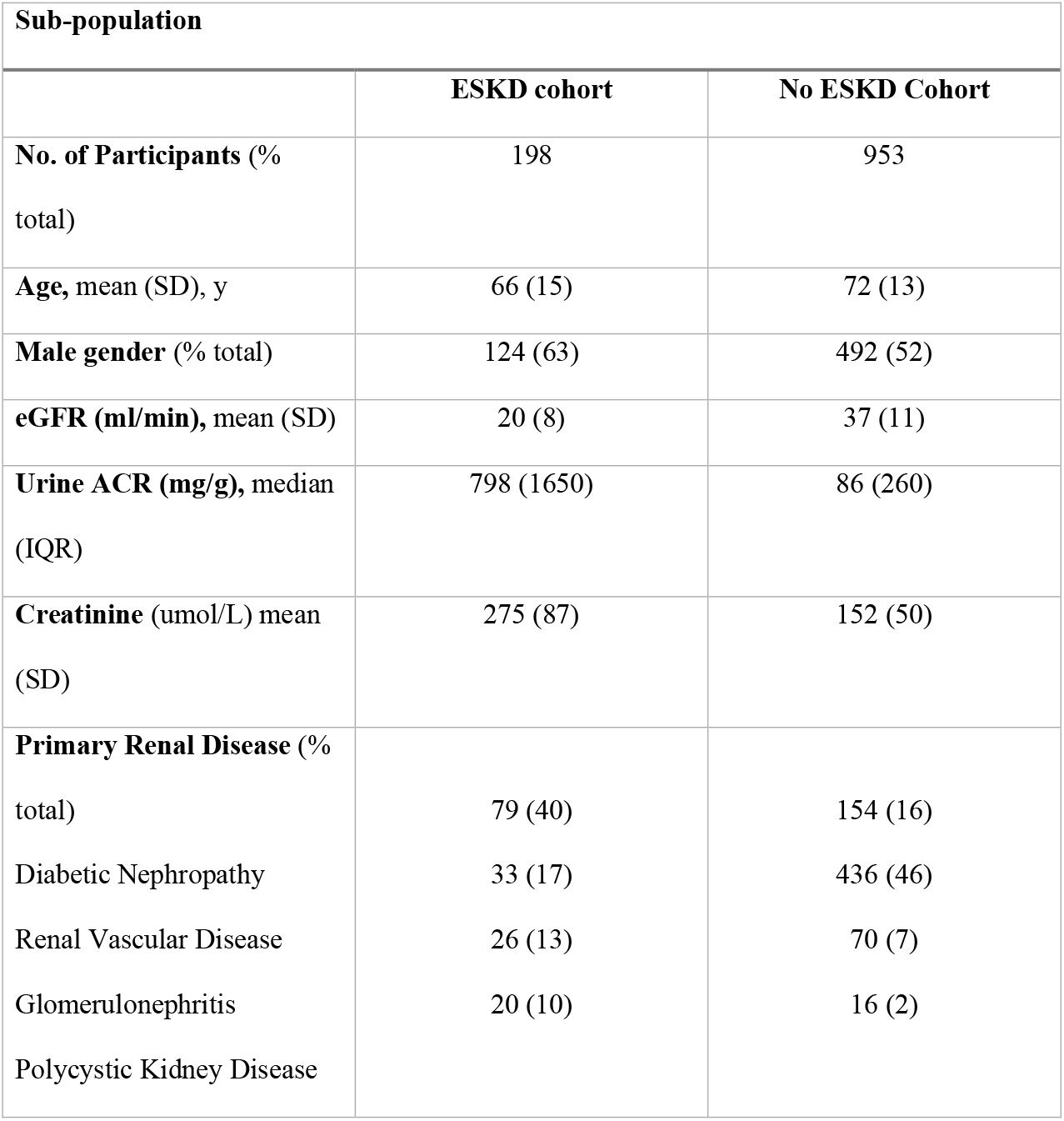

| Sub-population |  |  |
| --- | --- | --- |
|  | ESKD cohort | No ESKD Cohort |
| **No. of Participants** (%)<br>total) | 198 | 953 |
| **Age**, mean (SD), y | 66 (15) | 72 (13) |
| **Male gender** (%) total) | 124 (63) | 492 (52) |
| **eGFR (ml/min)**, mean (SD) | 20 (8) | 37 (11) |
| **Urine ACR (mg/g)**, median<br>(IQR) | 798 (1650) | 86 (260) |
| **Creatinine** (umol/L) mean<br>(SD) | 275 (87) | 152 (50) |
| **Primary Renal Disease** (%)<br>total) | 79 (40) | 154 (16) |
| Diabetic Nephropathy | 33 (17) | 436 (46) |
| Renal Vascular Disease | 26 (13) | 70 (7) |
| Glomerulonephritis | 20 (10) | 16 (2) |
| Polycystic Kidney Disease |  |  |

MNHHS = Metro North Hospital and Health District; SESLHD = South Eastern Sydney Local Health District; RRT = renal replacement therapy (including dialysis and transplant); eGFR = estimated glomerular filtration rate; \* age ≤ 18 or ≥ 90; \*\* incomplete 2 year pre-ESKD data (age, gender, ACR, eGFR); RF = random forest

