## Supplemental table 1 for "A comparison between a Random Forest model and the Kidney Failure Risk Equation to predict progression to kidney failure"

Appendix

sTable 1.

| Derived Population | | |
| --- | --- | --- |
|  | **KF cohort** | **No KF Cohort** |
| No. of Participants (% total) | 198 | 953 |
| Age, mean (SD), y | 66 (15) | 72 (13) |
| Male gender (% total) | 124 (63) | 492 (52) |
| eGFR (ml/min), mean (SD) | 20 (8) | 37 (11) |
| Urine ACR (mg/g), median (IQR) | 798 (1650) | 86 (260) |
| Creatinine (umol/L) mean (SD) | 275 (87) | 152 (50) |
| Primary Renal Disease (% total)  Diabetic Nephropathy  Renal Vascular Disease  Glomerulonephritis  Polycystic Kidney Disease | 79 (40)  33 (17)  26 (13)  20 (10) | 154 (16)  436 (46)  70 (7)  16 (2) |

KF = Kidney failure
